# The hospital burden of critical illness across global settings: a point-prevalence and cohort study in Malawi, Sri Lanka and Sweden

**DOI:** 10.1101/2024.03.14.24304275

**Authors:** Carl Otto Schell, Raphael Kayambankadzanja, Abigail Beane, Andreas Wellhagen, Chamira Kodippily, Anna Hvarfner, Grace Banda-Katha, Nalayini Jegathesan, Christoffer Hintze, Wageesha Wijesiriwardana, Martin Gerdin Wärnberg, Mtisunge Kachingwe, Petronella Bjurling-Sjöberg, Annie Kalibwe Mkandawire, Hampus Sjöstedt, Surenthirakumaran Rajendra, Cecilia Stålsby Lundborg, Miklos Lipcsey, Lisa Kurland, Rashan Haniffa, Tim Baker

## Abstract

**Importance:** Large unmet needs of critical care have been identified globally, but evidence to guide policy priorities is scarce. Available studies into the burden of critical illness have important limitations.

**Objective:** To assess the adult burden of critical illness in hospitals across global settings.

**Design, Setting, and Participants:** This was a prospective, observational, international, hospital-based, point-prevalence and cohort study in Malawi, Sri Lanka, and Sweden. On specific days, all adult in-patients in the eight study hospitals were examined for the presence of critical illness and followed up for hospital mortality.

**Exposure:** Patients with one or more severely deranged vital sign were classified as critically ill.

**Main Outcomes and Measures:** The primary study outcomes were the point-prevalence of critical illness and 30-day in-hospital mortality. In addition, we assessed the proportion of critically ill patients who were cared for in Intensive Care Units (ICU)s, and the association between critical illness and 30-day in-hospital mortality.

**Results:** Among 3652 hospitalized patients in countries of different socio-economic levels we found a point-prevalence of critical illness of 12.0% (95% CI, 11.0-13.1), with a hospital mortality of 18.7% (95% CI, 15.3-22.6). The odds ratio of death of critically ill compared to non-critically ill patients was 7.5 (95% CI, 5.4-10.2). Of the critically ill patients 3.9 % (95% CI, 2.4-6.1) were cared for in ICUs.

**Conclusions and Relevance:** The study has revealed a substantial burden of critical illness in hospitals from different global settings. One in eight hospital in-patients were critically ill, 19% of them died in hospital, and 96% of the critically ill patients were cared for outside ICUs. Implementing feasible, low-cost, critical care in general wards and units throughout hospitals would impact all critically ill patients and has potential to improve outcomes across all acute care specialties.

**Key Points:** *Question:* What is the burden of critical illness in hospitals in different global settings, and where are critically ill patients being cared for?

*Findings:* Among 3652 hospitalized patients in countries of different socio-economic levels (Malawi, Sri Lanka, and Sweden) we found a point-prevalence of critical illness of 12.0% (95% CI, 11.0-13.1), with a hospital mortality of 18.7% (95% CI, 15.3-22.6). The odds ratio of death of critically ill compared to non-critically ill patients was 7.5 (95% CI, 5.4-10.2). Of the critically ill patients 3.9 % (95% CI, 2.4-6.1) were cared for in Intensive Care Units (ICUs).

*Meaning:* Critical illness is common in hospitals and has a high mortality. Ensuring that feasible critical care interventions are implemented throughout hospitals including in general wards where more than nine in ten critically ill patients are cared for, has potential to improve outcomes across all medical specialties.

## Introduction

Critical illness is as a ‘state of ill health with vital organ dysfunction, a high risk of imminent death if care is not provided and the potential for reversibility.^1^ Regardless of underlying diagnosis, critically ill patients require similar initial actions to stabilize vital organ functions and prevent death. Such critical care interventions are needed wherever a critically ill patient is located. ^1,2^ Although many effective critical care interventions are low-cost and feasible throughout hospitals^3,4^ there is alarming evidence from different settings that they are frequently not provided. ^5-9^ Improving critical care has the potential to increase survival across medical disciplines. ^5,6,10-13^

The evidence to guide policy makers about critical illness and critical care is scarce. Decisions around priorities and investments in health care and research are often grounded in sources based on patients’ diagnoses where information about critical illness have not been captured.^14,15^ Most research into critical illness outcomes is confined to Intensive Care Units (ICUs), where advanced and high-cost critical care is provided. ICUs are sparce in rural and low resource settings where care needs are high.^13,16-18^ Per 100,000 population, ICU beds vary – from 0.1 in a Malawi, (low-income country) and 2.3 in Sri Lanka, (middle-income county) to 5.8 in Sweden and 35 in the USA (high-income countries).^19-21^

It has been estimated that the global incidence of critical illness among adults is 30-45 million per year, based on extrapolation from a North American ICU registry.^22^ This may be an underestimation as the adult incidence of sepsis alone is 24 million per year.^23^ Additionally, there are indications that critically ill patients may often be cared for outside ICUs.^24-30^ To guide policy making, there is a need for investigations that include critically ill patients throughout hospitals, across ward types, specialties, diagnoses, and socioeconomic levels. In this multi-centre global study, we aimed to assess the adult burden of critical illness in hospitals.

## Methods

### Study design and settings

This was a prospective, observational, international, hospital-based, point-prevalence and cohort study in Malawi, Sri Lanka, and Sweden. The study countries were chosen to include a low-income, middle-income and high-income country: the annual health expenditure (USD) per capita ranges from 33 in Malawi, to 151 in Sri Lanka and 6915 in Sweden.^31^ The study took place in eight public hospitals including first-line and referral hospitals in each country **(Table 1)**. Each hospital was assessed at least twice to control for seasonal variation. The principles of Good Clinical Practice were followed. Ethical permissions were provided in all settings - Malawi: College of Medicine (P.08/16/2007); Sri Lanka: University of Kelaniya (P/111/04/2018) and University of Jaffna (J/ERC/19/102/NDR/0205); Sweden: Ethical Review Board Stockholm (2017-1907-31-1).

**Table 1.** Country and hospital information.

| Country | Malawi |  | Sri Lanka |  | Sweden |  |  |  |
| --- | --- | --- | --- | --- | --- | --- | --- | --- |
| Population | 20,405,000 |  | 22,181,000 |  | 10,486,000 |  |  |  |
| GDP/capita (USD) | 645 |  | 3,354 |  | 56,424 |  |  |  |
| Life Expectancy (at birth) | 63 |  | 76 |  | 83 |  |  |  |
| Maternal mortality (per 100,000 live births) | 451 |  | 43 |  | 3 |  |  |  |
| Annual health expenditure per capita (USD) | 33 |  | 151 |  | 6915 |  |  |  |
| Hospital beds (per 100,000) | 130 |  | 420 |  | 210 |  |  |  |
| ICU beds per (100,000) | 0.1 |  | 2.1 |  | 5.7 |  |  |  |
| Hospital | Queen Elizabeth Central Hospital | Chiradzulu District Hospital | Monaragala District General Hospital | Point Pedro Base Hospital | Universitets-sjukhuset Örebro | Mälarsjukhuset Eskilstuna | Nyköpings Lasarett | Kullbergsska Sjukhuset |
| Hospital type | Government run teaching hospital | Government run first-level hospital | Government run first-level and referral hospital | Government run first-level hospital | Government run university hospital | Government run first-level and referral hospital | Government first-level hospital | Government first-level hospital |
| Immediate catchment population(n) | 1,000,000 | 240,000 |  | 150,000 | 200,000 | 146,000 | 96,000 | 59,000 |
| Extended catchment population(n) | 9,000,000 | 240,000 | 556,000 | 175,000 |  |  |  |  |
| Total Adult somatic Hospital beds (n) | 1150 | 240 | 460 | 221 | 402 | 254 | 122 | 57 |
| ICU Beds (n) | 8 | 0 | 12 | 0 |  | 7.7 | 6.5 |  |
| Admissions per year (n) | 40000 | 10000 |  |  |  |  |  |  |
| Adult somatic admissions per year (n) |  |  | 48000 | 24448 |  | 18821 | 10 823 | 4 250 |
| Mean length of stay |  | 2 days |  |  |  |  |  |  |
| Mean length of stay adult somatic |  |  | 2 days | 2 days |  | 4.6 | 4.2 | 4.6 |
| Hospital mortality | 7% | 15% |  | 0.6% |  | 3,6% | 3,1% | 3,9% |

### Participants and outcomes

In each hospital on the days of point-prevalence assessment, all patients in all somatic wards above 18 years of age were included in the study. Participants who were able to, provided informed consent. For the validity of the study, we included patients with reduced consciousness in absence of objection from the patient (verbal or non-verbal) or from the next of kin. We could not include patients who were in operating theaters or were absent and could not be found later in the day. The study excluded women in active labor, patients who were not admitted to hospital (neither had stayed, or planned to stay overnight), and moribund patients identified as “*dying”* by the attending nurse. All participants had their vital signs examined for presence of critical illness, and they were followed up for hospital mortality, censored at 30 days. We used the term *burden* of critical illness when referring to the impact of critical illness - both occurrence (prevalence) and consequence (mortality). The primary study outcomes were the point-prevalence of critical illness and the 30-day in-hospital mortality of patients with critical illness. In addition, we assessed the proportion of critically ill patients who were cared for in ICUs, and the association between critical illness and 30- day in-hospital mortality.

### Variables

Critical illness was defined as “*a state of ill health with vital organ dysfunction, a high risk of imminent death if care is not provided and a potential for reversibility”* ^1^ and operationalized by classifying a critically ill patient as someone with one or more severely deranged vital sign at the point prevalence examination. Such criteria are independent of ward type and specialty and are pragmatic for use in clinical practice. The *a priori* decided cutoffs for critical illness are based on triggers for clinical intervention used at Karolinska University Hospital (Sweden) and in Tanzania **(Table 2)**.^27,30,32,33^

**Table 2.** Parameters and cutoffs for danger signs.

|  |  | Danger Signs |
| --- | --- | --- |
| **A** | **Airway sounds** | Stridor or Gurgling or Snoring |
| **B** | **Respiratory rate (per minute)**<br>**Oxygen saturation (%)** | <8 or >30<br><90 |
| **C** | **Heart rate (per minute)**<br>**Systolic blood pressure (mmHg)** | <40 or >130<br><90 |
| **D** | **Conscious level** | Glasgow Coma Scale <9 |

The patient’s hospital records were used for clinical information about age, sex, diagnosis, specialty, and decision to not resuscitate in case of cardiac arrest (DNR). ICU-beds were classified per hospital definition. All other patients were classified as located in general wards. Some hospitals described some ward beds as providing a higher care intensity, “high care beds”, or “high dependency unit” beds. However, the care and interventions available in such locations varied substantially between settings and precluded a formal analysis.

### Data collection

Data collections took place on individual days between 2017 and 2019 in the study hospitals. All hospital wards and units were visited, regardless of admitting specialty. Teams of nurses and students of health care professions went from ward to ward to include all the hospital in-patients and assess their vital signs. A senior health worker or researcher supervised in each ward to ensure quality data collection. Prior to this, the data collectors had a day of practice and training on research methods, ethics, study methods, equipment usage, and standardized vital signs assessments. The equipment was quality tested before each data collection and included automatic blood-pressure monitors, pulse oximeters, thermometers, and clocks. Abnormal vital signs were re-checked, and alternative methods were used if a vital sign could not be assessed (e.g. using manual blood pressure measurement). The nurse-in-charge of the ward was notified immediately when a patient was identified as critically ill. The research team offered to document all vital signs collected in the clinical records for use in patient care.

In Malawi and Sri Lanka, clinical information was extracted from the paper-based patient records and outcomes were collected through follow-up of records and hospital administrative data. In Sweden, these data were collected from electronic medical records.

### Statistical methods

We used percentages to present point-prevalence, in-hospital mortality, and location of the critically ill. The association of critical illness and in-hospital mortality was assessed using odds ratios (OR) in crude logistic regression models and adjusted odds ratios (aOR) in models including prespecified potential confounders: for age, sex and country. Exact logistic regression was used for analyse at country level. Missing data for a single vital sign were classified as not being a danger sign (the most common value), enabling use of the other vital signs to classify the patient’s critical illness status.

Participants who were lost to follow up or had missing data for age, sex or country were excluded from analyses. A 95% confidence interval was used in reporting findings. Stata/IC 15 (Stata Corp, College Station, TX) was used for analyses.

## Results

A total of 3682 participants were initially included. A final cohort of 3652 participants was used for analyses, after exclusion of twenty patients who were lost to follow-up (18 from Malawi and 2 from Sri Lanka) and ten patients from Malawi with missing data for age. Out of 21912 expected data points for vital signs (6 per participant), 72 (0.03%) were missing.

Women comprised 2015 (55%) of all participants and 224 (51%) of the critically ill. The median age of the cohort was 58 years [IQR34-75] and 61 years [IQR 37-76] among critically ill patients. A majority of patients were admitted to a medical department: 1846 (51%) of all patients and 327(74%) of the critically ill.

In the study countries, there were 653 (59%) women in Malawi, 436 (60%) in Sri Lanka, and 926 (51%) in Sweden. The median age was 35 years [IQR 26-49] in Malawi, 41 years [IQR 30-62] in Sri Lanka and 73 years [IQR61-82] Sweden. There were two patients having a DNR in Malawi (0.2%) none in Sri Lanka and 346 in Sweden (19%). Clinical characteristics of the cohort are presented in **Table 3**.

**Table 3.** Participant characteristics.

|  | ALL |  | MALAWI |  | SRI LANKA |  | SWEDEN |  |
| --- | --- | --- | --- | --- | --- | --- | --- | --- |
|  | All | Critical | All | Critical | All | Critical | All | Critical |
| Participants, n | 3652 | 439 | 1107 | 204 | 723 | 43 | 1822 | 192 |
| Death in hospital, n (%) | 178(4.9%) | 82(19%) | 85(7.1%) | 42(21%) | 8(1.1%) | 6(14%) | 85(4.7%) | 34(18%) |
| Female, n (%) | 2015(55%) | 224(51%) | 653(59%) | 119(58%) | 436(60%) | 17(40%) | 926(51%) | 88(46%) |
| Age, years [IQR] | 58[34-75] | 61[3-76] | 35[26-49] | 38[30-51] | 41[30-62] | 56[39-68] | 73[61-82] | 76[69-86] |
| Specialty, n (% per column) |  |  |  |  |  |  |  |  |
| - Medicine | 1846(51%) | 327(74%) | 461(42%) | 143(70%) | 244(34%) | 24(56%) | 1141(62%) | 160(83%) |
| - OBG | 653(18%) | 24(5.5%) | 283(26%) | 16(7.8%) | 260(36%) | 7(16%) | 110(6.0%) | 1(0.5%) |
| - Surgery | 1151(31%) | 87(20%) | 362(33%) | 45(22%) | 218(30%) | 11(26%) | 571(31%) | 31(16.2%) |
| - Unknown | 2 | 1 | 1 | - | 1 | 1(2.3%) | - | - |
| (DNR), n (%) | 348(10%) | 81(18%) | 2(0.2%) | 2(1%) | 0 | 0 | 346(19%) | 79(41%) |
| Level of care, n (% per column) |  |  |  |  |  |  |  |  |
| - ICU | 52(1.4%) | 17 (3.9%) | 5(0.5%) | 3(1.5%) | 7(1.0%) | 3(7.0%) | 40(2.2%) | 11(5.7%) |
| - General Ward | 3600(99%) | 422(96%) | 1102(99%) | 201(98.5%) | 716(99%) | 40(93%) | 1782(98%) | 181(94%) |

DNR indicates Decision to Not Resuscitate in case of cardiac arrest; ICU Intensive Care Unit; IQR indicates Inter Quartile Range

Critical illness was present in 439 patients, corresponding to a point-prevalence of critical illness of 12.0% (95% CI 11.0-13.1). The critically ill patients had a hospital mortality of 18.7% (15.5-22.8). Of the critically ill patients, 17 (3.8% (2.4-6.0), were cared for in an ICU and 422 (96%) in a general ward. Outcome data are presented in **Table 4**.

**Table 4.** Critical illness: point-prevalence, mortality, and proportion in ICU.

|  | All | Malawi | Sri Lanka | Sweden |
| --- | --- | --- | --- | --- |
| Prevalence Critical Illness %, (95% CI) | 12.0 (11.0-13.1) | 18.4(16.3-20.1) | 5.9 (4.4-7.9) | 10.5 (9.2-12.0) |
| Mortality Critical Illness %, (95% CI) | 18.7(15.3-22.6) | 20.6(15.6-26.7) | 14.0(6.3-28.3) | 17.8(12.9-23.8) |
| Proportion of critically ill patients in ICU %, (95% CI) | 3.9 (2.4-6.1) | 1.5 (0.5-4.5) | 7.0 (2.2-20.1) | 5.7 (3.2-10.1) |

CI indicates confidence interval; ICU Intensive Care Unit

In the whole cohort, the association between critical illness and death was OR 7.5 (5.4-10.2). In the model adjusted for age and sex, aOR was 7.3 (5.3-10.0). In the model adjusted for age, sex and country, aOR was 6.1 (4.4-8.4) The use of cubic splines to ensure that the association between age and death was not underestimated did not increase the association in any model and so was not used. **(Table 5)**

**Table 5.**
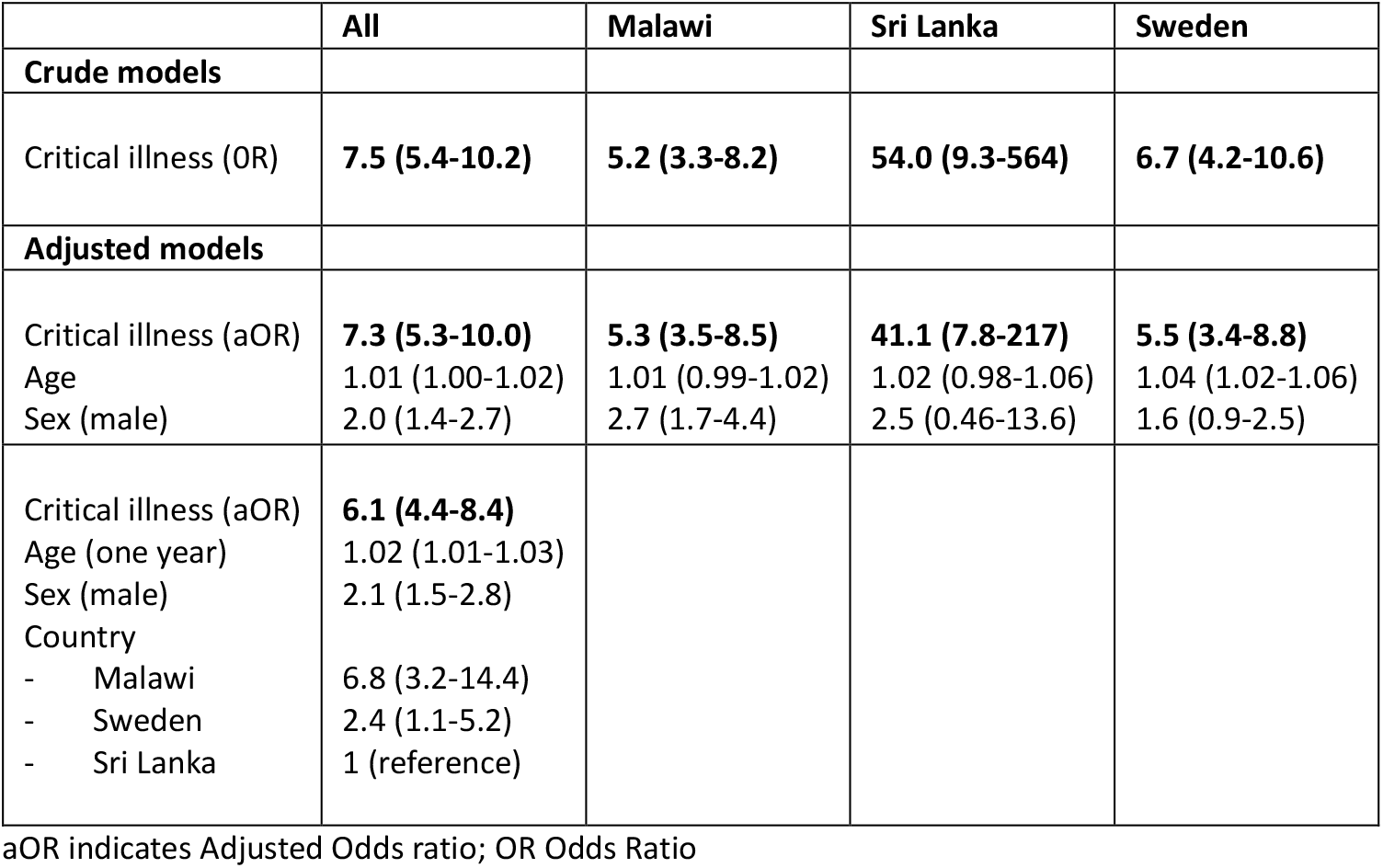
The association between critical illness and 30-day in-hospital mortality.

|  | All | Malawi | Sri Lanka | Sweden |
| --- | --- | --- | --- | --- |
| **Crude models** |  |  |  |  |
| Critical illness (OR) | **7.5 (5.4-10.2)** | **5.2 (3.3-8.2)** | **54.0 (9.3-564)** | **6.7 (4.2-10.6)** |
| **Adjusted models** |  |  |  |  |
| Critical illness (aOR) | **7.3 (5.3-10.0)** | **5.3 (3.5-8.5)** | **41.1 (7.8-217)** | **5.5 (3.4-8.8)** |
| Age | 1.01 (1.00-1.02) | 1.01 (0.99-1.02) | 1.02 (0.98-1.06) | 1.04 (1.02-1.06) |
| Sex (male) | 2.0 (1.4-2.7) | 2.7 (1.7-4.4) | 2.5 (0.46-13.6) | 1.6 (0.9-2.5) |
| Critical illness (aOR) | **6.1 (4.4-8.4)** |  |  |  |
| Age (one year) | 1.02 (1.01-1.03) |  |  |  |
| Sex (male) | 2.1 (1.5-2.8) |  |  |  |
| Country |  |  |  |  |
| - Malawi | 6.8 (3.2-14.4) |  |  |  |
| - Sweden | 2.4 (1.1-5.2) |  |  |  |
| - Sri Lanka | 1 (reference) |  |  |  |

aOR indicates Adjusted Odds ratio; OR Odds Ratio

## Discussion

In this prospective point prevalence and cohort study of all in-patients in eight hospitals from Malawi, Sri Lanka and Sweden, we found a substantial burden of critical illness. The point-prevalence of critical illness was 12% and the critically ill had a hospital mortality of 19%. Critically ill patients had a significantly higher odds of in-hospital mortality than non-critically (OR 7.5) and of the critically ill, 96% were cared for outside ICUs.

The point-prevalence found is consistent with data that could be extracted from previous single-centre studies with other aims. Among hospital patients in Finland and Sweden, 8% and 12-14% respectively had a severely deranged vital sign ^25-27^. In medical and surgical wards in Uganda, 12% of patients had a “critical” modified early waring score of more than 5. ^28^ Our results support previous indirect estimations of a substantial global burden of critical illness.^22^

The mortality of critically ill patients in our study is high compared to other patient groups and diagnoses of public interest. For example, patients admitted for care in Swedish ICUs had a hospital mortality of 14%.^34^ Among patients with COVID-19 during the first wave in USA, 20% died in hospital.^35^ Acute myocardial infarction with ST-elevation had 30-day mortalities of 2.4-5.0%, 1.0% and 0.4% in Sub-Saharan Africa, South Asia and North Europe respectively.^28-30^ Our findings confirm that critical illness, as identified in a pragmatic way using deranged vital signs is a high-risk condition with very high rates of mortality.

Most critically ill patients are cared for outside ICUs, and this is not limited to low resource settings. As countries have such large differences in the number of ICU beds – 350 times more beds per 100,000 population in the USA than in Malawi^21^ – the presence of critically ill patients in general wards might be thought to be specific to low-income countries. This is neither supported by our results nor by previous research.^25-27,29^

There are likely explanations behind specific findings in each of the study countries. Sri Lanka, the middle-income country, had the lowest critical illness prevalence and lowest critical-illness mortality but also the lowest mortality of non-critically ill patients (0.3%). One explanation for this may be a higher number of hospital beds (420) per 100,000 population than Sweden (210) and Malawi (130). This could lead to a lower threshold for admitting patients to hospital in Sri Lanka than the other countries, thus “diluting” the proportion of critically ill patients.^31^ Finding the highest critical illness mortality in Malawi (21%), was expected, as resources are far scarcer than in the other countries, affecting the determinants of health and the resources available for health care.^31^ In low-income countries, critical illness outside hospital may also be common, since limited access may delay or preclude care.^36^ The high mortality of critically ill patients in Sweden of 18% was an interesting finding, and is likely explained by the high median age of the patients (73 years), above which frailty and multimorbidity are common.^37^

### Implications

The findings suggest a need for health systems to recognize and prioritize critical illness throughout hospitals. This would not be an unachievable goal. In fact the unmet needs of care for most critically ill patients ^5-9^ can be provided outside ICUs.^4,11,13,38^ The recently defined Essential Emergency and Critical Care (EECC) includes 40 foundational interventions selected for clinical effectiveness and feasibility in all hospital settings, such as triage, airway protection and oxygen therapy.^3,12^ EECC aligns with the WHO’s Fair Priorities framework to maximize the population impact of care interventions.^39^ Ward-based critical care has lower costs and has been shown to be more cost-effective than ICU care for many patients groups ^40-43^. In Tanzania, EECC has been estimated to be highly cost-effective at 14 USD per healthy life-year gained.^44^ Ensuring EECC is provided to all patients who need it throughout hospitals and health systems would seem to be the rational first step when improving critical care services.^11^ In cases where the fundamental critical care is not enough to stabilize organ functions, high dependency units (HDUs) may be a reasonable subsequent step and may be more equitable and effective than an expansion of resource-intensive ICUs.^45,46^ Governments that use a strategy to improve critical care by starting from the fundamental level could reach all critically ill patients, be cost-effective, and have impact at population level.^10,11,47^

### Strengths and limitations

The prospective examination of all in-patients in this study, regardless of diagnosis and location in hospital, minimized the risk for selection bias and misclassification. The quality of the data collection increased internal validity through high inclusion rates, few missing data points, and accuracy of the data. Studying hospitals in a low-, a middle- and a high-income country enabled the inclusion of patients from settings with a large global variation. The feasible clinical criteria for the identification of critical illness and the pragmatic data collection methods that were used enable replication in health facility audits and larger studies.

There are limitations. First, the pragmatic criteria for critical illness may have missed high-risk patients whose vital signs were insufficiently deranged or who had been stabilized by healthcare interventions. Conversely, some patients with adapted physiology due to chronic disease may have been misclassified as critically ill. Second, we could not include patients in operating theatres, some of whom may have been critically ill. Third, data were collected during working hours and the burden of critical illness may be altered during weekends and nights.^48^ Fourth, the ethical imperative to inform the ward nurses about the patients who were critically ill may have led to improved care and reduced critical illness mortality in the cohort. Last, the number of countries and hospitals included limit generalization to all other settings, but we do not have reasons to think that the burden of critical illness would be markedly different in other hospitals.

## Conclusion

The study has revealed a substantial burden of critical illness in hospitals from different global settings. One in eight hospital in-patients were critically ill, 19% died in hospital, and 96% of the critically ill patients were cared for outside ICUs. Implementing feasible, low-cost, critical care in general wards and units throughout hospitals would impact all critically ill patients and has potential to improve outcomes across all acute care specialties.

## Data Availability

Due to the sensitive nature of the information about severely unwell patients, and the requirements of the College of Medicine Research and Ethics Committee, data from the study are not publicly available. Data can be requested by contacting the corresponding author including a reasonable motivation for the request for the data and a study plan.

## ABBREVIATIONS

aOR: Adjusted Odds Ratio
DNR: Decision to Not Resuscitate in case of cardiac arrest
EECC: Essential Emergency and Critical Care
HDU: High Dependency Unit
ICU: Intensive Care Unit
IQR: Inter Quartile Range
OR: Odds Ratio
USA: United States of America
USD: United States Dollar

